# Disparities in self-reported Mental Health, Physical Health, and Substance Use Across Sexual Orientations in Canada

**DOI:** 10.1101/2024.05.24.24307865

**Authors:** Zachary Bellows, Chungah Kim, Yihong Bai, Peiya Cao, Antony Chum

## Abstract

**Background:** While prior studies have shown LGB individuals have elevated risk of poor mental health, poor physical health, and substance use, existing study designs may be improved by using representative samples, wider ranges of health outcomes, heterosexual comparison groups, and disaggregated data. The goal of this study is to provide estimates of multiple health disparities across sexual orientations in Canada based on these principles.

**Methods:** Using data from 2009-2014 Canadian Community Health Surveys, a sample of 19,980,000 weighted individuals was created. Outcomes included mental health, physical health, binge drinking, illicit drug use, and cannabis use. The study used logistic regression models adjusted by covariates, stratified by sex, to estimate health disparities across sexual orientations.

**Results:** Among LGB individuals, there was evidence for elevated risk of poor mental health (i.e. gay men, bisexual men, bisexual women), poor physical health (i.e. bisexual men, bisexual women), binge drinking (i.e. lesbians, bisexual women), illicit drug use (i.e. lesbians, bisexual women), and cannabis use (i.e. lesbians, bisexual women) relative to their heterosexual counterparts. Those identifying as ‘don’t know’ or ‘refuse’ showed reduced odds of substance use. Bisexual women exhibited highest disparities in health outcomes, e.g. OR=3.3, 95% 2.58 to 4.22 for poor mental health. Trends over time showed worsening mental health among bisexual women (relative to changes in heterosexual women), and decreasing substance use in gay and bisexual men, and lesbians.

**Conclusion:** This study highlights health disparities across sexual orientations in Canada, especially bisexual women, calling for targeted interventions (e.g. increased training of service providers in working with bisexual women and community outreach against biphobia). Future research should aim to explore these disparities longitudinally while also including the use of administrative-linked health data to reduce potential bias in self-reported data.

## Introduction

Promoting health equity in marginalised communities is a fundamental objective of public health; however, to address these disparities effectively, it is crucial to understand their magnitude. The purpose of this study is to provide an estimate of health disparities across sexual orientations with regards to mental health, physical health, alcohol consumption, and substance use (illicit drug use and cannabis use).

### Prior literature and the minority stress model

The minority stress model posits that belonging to a minority group inherently presents unique social challenges, leading to elevated stress responses and adverse health outcomes[1,2]. With regards to health disparities across sexual orientation, the model specifically highlights how clashes with heteronormative culture can engender environments where sexual minorities face distinctive stressors such as internalisation of negative societal attitudes, anticipation of rejection, and experiences of discrimination[3]. These stressors subsequently contribute to poor mental health [4][5], physical health conditions [6][7], and may even develop into substance misuse as a coping strategy [8][9].

While the minority stress model has contributed to the theoretical foundations for the study of health disparities across sexual orientations, prior studies are characterised by key limitations that we aim to address in our present study, they include: 1) testing the minority stress theory with a narrow set of health outcomes, 2) limitations of convenience samples, 3) missing heterosexual comparison group, and 4) the lack of appropriate disaggregation across sexual orientation groups.

### Testing the minority stress theory using a narrow set of health outcomes

White et al. argue the traditional explanatory models used in epidemiology studies are often “disease specific”, identifying risk factors for specific health conditions (1); however, social exposures (e.g. the impact of discrimination) may have a more generalised and cumulative health impact rather than being specific to one illness, and the focus on a single disease/condition may not adequately capture the generalised impact of the social exposure on overall health[10]. The practice of disease-specific epidemiological investigations is also common among studies of sexual minority health, with many prior studies examining disparities across sexual orientation in a narrow range of health outcomes such as substance use[11], mental health[12], and physical health[13]. Yet, the mechanism through which minority stress is theorised to impact health outcomes, namely minority stress [8], are not specific to any given disorder or condition and are expected to have a generalised, cumulative health impact. Therefore, studies that focus narrowly on a single disease/disorder, rather than focusing on domains of health (such as mental health or physical health), have been argued by Frost [14], to risk “false null” findings that could imply social disadvantage does not affect the health of sexual minorities. Therefore, if possible, studies of disparities in health status across sexual orientation would benefit from the examination of a wide range of health outcomes to capture the generalised health impact of minority stress.

### Limitations of convenience samples

A number of studies that examined sexual minority health have relied on convenience samples or small, non-representative samples[12,15–18]. Convenience sampling involves selecting participants based on their availability and ease of recruitment[19], which limits the generalizability of the findings since the characteristics of those who volunteer as participants may systematically differ from non-participants. Among these studies, participants were often recruited at Pride and community events, with postcards advertising the study being distributed LGBTQ+ (i.e. lesbian, gay, bisexual, transgender, queer, and others) community organisations, and was supplemented by advertisement on radio, social media websites, and mobile meet-up apps such as Grindr. These recruitment strategies may lead to a sample who are over-represented by those who are well connected to the LGBTQ+ community, while those who are isolated from the LGBTQ+ community or not out of the closet are less likely to be recruited leading to potential sampling bias[19]. Therefore, if people who are isolated have a systematically different health status compared to those who are well-connected, as suggested by prior literature[20], then prior studies using convenience samples will likely be biased.

### Missing heterosexual comparison group

Some previous studies have investigated sexual minority health without including a heterosexual comparison group [12,15,21,22][23] [24] [12,15,21,22], which makes it difficult to quantify disparities and measure the impact of minority stress, since comparing health outcomes between sexual minority groups who face minority stress and heterosexual individuals are required to highlight disparity. Using a heterosexual comparison group provides a reference point that allows us to determine the extent of health disparities across sexual orientations. While these studies helped characterise health problems in sexual minority individuals, the use of comparison groups would help contextualise these health problems (i.e. understand the risk irrespective of sexual minority status) and shed light on the specific challenges faced by lesbian, gay, and bisexual (LGB) individuals.

### Issues with no disaggregation across sex or sexual minority groups

Another limitation in the literature is common practice of the aggregating lesbian, gay, and bisexual, individuals into a single “LGB” or sexual minority category in prior studies [25–27][28]. This approach can be problematic since it may obscure important disparities between these groups. Specifically, bisexual individuals may face unique challenges: 1) marginalisation by both heterosexual and gay/lesbian communities[29], 2) experience of biphobia and invalidation (e.g. being dismissed or excluded based on prejudice and stereotypes about bisexual individuals)[30], and 3) encounter mistrust and stereotypes (e.g. related to their ability to commit) that affect their relationships and overall well being[31]. Disaggregating gay/lesbians from bisexuals may reveal important health disparities that are hidden in the data.

Another related issue is that even when a study disaggregates gays/lesbians from bisexuals, men and women may be aggregated into a single group, and gender/sex is typically used as an adjustment variable in these studies[32–34], which can lead to misestimations since there may be significant gender/sex differences within a given sexual orientation (e.g. gay men vs. lesbians). While disaggregated analysis by sex allows second level sex differences to be revealed across all model covariates[35], it comes at a cost to statistical power that may be too high for many studies except for large scale population-based samples collected by national statistical bureaus.

Our study objectives include 1) investigating the disparity in mental health, physical health, binge drinking, illicit drug use, and cannabis use across sexual orientations; and 2) exploring the trends in disparities over time between 2009-14. Unlike earlier studies that focused on limited health outcomes, our research encompasses mental health, physical health, binge drinking, illicit drug use, and cannabis use, providing a comprehensive overview of the health challenges faced by sexual minorities. We utilize data from a nationally representative survey, including sexual minorities from remote areas and those disconnected from LGBTQ+ communities. Additionally, unlike some previous studies, we include a heterosexual comparison group to assess the impact of minority stress. We also disaggregate data by sexual orientation and sex, offering insights into the diverse experiences within the sexual minority population.

## Methods

### Sample

The Canadian Community Health Survey (CCHS) is a cross-sectional survey conducted annually across Canada, utilizing multistage sampling and computer-assisted telephone interviews in English or French. This study includes data from six cycles (2009-2014), comprising a weighted sample of 19,980,000 individuals aged 18-59. Survey weights were used to ensure the representativeness of the Canadian population. The response rates of the six cycles ranged from 87.3% [36] to 89.3% [37].

Secondary data analysis was conducted without obtaining written informed consent from participants for this research. This study adhered to the data confidentiality guidelines set by Statistics Canada and the Statistics Canada Research Data Centre. The data were anonymized, and the research team did not have access to the personal identifiers of CCHS participants. The study has received ethical approval from the York University Research Ethics Board (certificate number: 20-134-CHUM).

### Exposure variable

Participants were asked to indicate their sex as either male or female. In order to determine sexual orientation, participants were asked, “Do you consider yourself to be…”, and were given the following response options: heterosexual, homosexual, bisexual, or ‘don’t know’, or ‘refuse to say’. When compared to questionnaires that encompass sexual identity, behaviour, and attraction through a multi-question approach, this single-question approach has been shown to be a reliable measure and has displayed a strong correlation with sexual identity (kappa statistic of 0.89)[38]. In previous versions of the CCHS, the single-question method successfully identified 99.3% of participants who identified as a sexual minority based on a comprehensive questionnaire, as well as 84.2% of those who reported engaging in same-sex relationships at any point in their lives[38]. In this study, sexual orientation is represented by five categories: heterosexual, gay/lesbian (homosexual), bisexual, ‘don’t know’, and ‘refuse to say’.

### Outcome Variables

Outcomes included mental and physical health, binge drinking, illicit drug use, and cannabis use. Mental health and physical health were dichotomized as: excellent, very good, good, vs. fair, poor for physical and mental health. A scoping review on the validity of single item self-rated mental health found that ratings of fair and poor self-rated mental health had 4.57 to 9.97 times higher risk of being diagnosed with major depressive disorder[39]. Similarly, fair and poor physical self-rated health has been associated with a 2-fold higher mortality risk compared with persons reporting a higher health status[40].

Binge drinking was determined by any instance of self-reported binge drinking in the past 12 months, where binge drinking is defined by the CCHS as a male having more than four standard alcoholic drinks in one occasion or a female having more than three standard alcoholic drinks in one occasion. Illicit drug use was determined by a derived drug use variable based on questions asking the participant if they used the following drugs in the past 12 months: cocaine or crack, speed/amphetamines, ecstasy/MDMA, hallucinogens, PCP (Phencyclidine), LSD/acid (Lysergic acid diethylamide), sniffing glue, gasoline, other solvents, and heroin. Cannabis use was determined by its use in the past 12 months, based on a self-reported response to essentially: “During the past 12 months have you used marijuana?”. Previous studies have provided validation for self-administered single-item screening questions aimed at detecting unhealthy substance use. These studies indicate that individuals with substance use conditions are at least three times more likely to yield a positive screen and are less than one-third as likely to yield a negative screen [41]. Furthermore, the reliability of self-rated substance consumption measures has been demonstrated in prior literature. For instance, the reliability coefficient (kappa) for binge drinking alcohol stands at 0.76, while it ranges from 0.72 (for hallucinogens) to 0.76 (for cocaine) for illicit drug use. Additionally, the reliability coefficient for cannabis use is reported at 0.82 [42]. These findings underscore the utility and validity of self-administered screening questions and self-rated substance consumption measures in identifying and assessing unhealthy substance use behaviours.

### Covariates

The following variables were used in the models as statistical controls: 1) year of birth (continuous), 2) marital status (married or common law vs. single, separated, divorced, or widowed), 3) educational attainment (achieved at least a post-secondary degree/certificate vs. not), 4) student status (current student vs. not), 5) self-reported ethnic minority status (yes vs. no), 6) employment status (Any work in the last year vs. not), 7) rurality status (yes vs. no) based on prescribed rural postal codes[43], 8) province of residence, 9) year of interview as dummies (2009-2014), and 10) income.

### Statistical Analysis

First, cross-tabulations of sexual orientations and outcomes, along with covariates, are calculated. Second, logistic regression models were used to estimate differences in outcomes across sexual orientations. All models were stratified by sex. Unadjusted models and adjusted models were estimated for each outcome. Multiple imputation by chained equations was used to impute missing values in all covariates.

The following sensitivity tests were also performed: 1) models using a modified Poisson regression; 2) models using complete case analyses (dropping cases with missing data instead of multiple imputations); and 3) models with interactions between sexual orientation and year of interview to check for potential change over the study period. All analysis was conducted using STATA v17.

## Results

Table 1 shows the descriptive statistics of the study participants (weighted n= 19,980,000), which includes cross-tabulations between sexual orientation with each outcome and sociodemographic covariates (e.g. age, income, ethnicity, etc.). Bisexual respondents reported the highest rates of poor physical (17.67%), mental health (20.47%), binge drinking (61.4%), illicit drug use (13.02%), and cannabis use (13.02%). Bisexuals and ‘don’t know’ respondents tended to occupy lower income quintiles, with bisexuals notably concentrated in the lowest quintile (30.23%). Homosexuals and heterosexuals appeared more frequently in higher income quintiles. Homosexual and bisexual groups were less likely to be married and living in rural areas. Employment rates were highest among heterosexual and homosexual respondents.

**Table 1:** Descriptive Statistics by Sexual Orientation: Mental and Physical Health, Substance Use, Demographics, and Socioeconomic Indicators from the Canadian Community Health Survey.

|  | Heterosexual | Homosexual | Bisexual | Don't Know | Refuse | Total |
| --- | --- | --- | --- | --- | --- | --- |
| **Number of Persons** | 19,122,000 | 284,000 | 215,000 | 59,000 | 120,000 | 19,980,000 |
| **Poor Physical Health, N (column %)** | 1,623,000 (8.49%) | 28,000 (9.86%) | 38,000 (17.67%) | 8,000 (13.56%) | 18,000 (15.00%) | 1,767,000 (8.84%) |
| **Poor Mental Health, N (column %)** | 1,121,000 (5.86%) | 26,000 (9.15%) | 44,000 (20.47%) | 6,000 (10.17%) | 13,000 (10.83%) | 1,210,000 (6.06%) |
| **Binge Drank Alcohol<sup>1</sup>, mean (column %)** | 9,527,000 (49.82%) | 169,000 (59.51%) | 132,000 (61.40%) | 11,000 (18.64%) | 29,000 (24.17%) | 9,906,000 (49.58%) |
| **Illicit Drug Use<sup>2</sup>, N (column %)** | 962,000 (5.03%) | 22,000 (7.75%) | 28,000 (13.02%) | 1,000 (1.69%) | 3,000 (2.50%) | 1,016,000 (5.09%) |
| **Cannabis Use<sup>3</sup>, N (column %)** | 987,000 (5.16%) | 22,000 (7.75%) | 28,000 (13.02%) | 1,000 (1.69%) | 3,000 (2.50%) | 1,041,000 (5.21%) |
| **Mean Age (standard deviation)** | 39 (12.11) | 39 (12.36) | 32 (11.96) | 41 (12.41) | 40 (12.39) | 39 (12.15) |
| **Married, N (column %)** | 11,948,000 (62.48%) | 102,000 (35.92%) | 64,000 (29.77%) | 31,000 (52.54%) | 58,000 (48.33%) | 12,280,000 (61.46%) |
| **Has Completed Post-Secondary, N (column %)** | 12,328,000 (64.47%) | 208,000 (73.24%) | 104,000 (48.37%) | 26,000 (44.07%) | 66,000 (55.00%) | 12,783,000 (63.98%) |
| **Current Student, N (column %)** | 2,577,000 (13.48%) | 46,000 (16.20%) | 55,000 (25.58%) | 6,000 (10.17%) | 17,000 (14.17%) | 2,727,000 (13.65%) |
| **White, N (column %)** | 14,460,000 (75.62%) | 240,000 (84.51%) | 164,000 (76.23%) | 30,000 (50.85%) | 62,000 (51.67%) | 15,082,000 (75.49%) |
| **Employed in the last year, N (column %)** | 16,767,000 (87.68%) | 255,000 (89.79%) | 173,000 (80.47%) | 39,000 (66.10%) | 90,000 (75.00%) | 17,324,000 (86.71%) |
| **Lives Rural, N (column %)** | 2,898,000 (15.16%) | 26,000 (9.15%) | 22,000 (10.23%) | 8,000 (13.56%) | 11,000 (9.17%) | 2,994,000 (14.98%) |
| **Income Quintile** |  |  |  |  |  |  |
| **Q1 (Lowest), N (column %)** | 3,082,000 (16.11%) | 42,000 (14.79%) | 65,000 (30.23%) | 21,000 (35.59%) | 35,000 (29.17%) | 3,291,000 (16.47%) |
| **Q2, N (column %)** | 3,211,000 (16.79%) | 45,000 (15.85%) | 41,000 (19.07%) | 9,000 (15.25%) | 20,000 (16.67%) | 3,366,000 (16.85%) |
| **Q3, N (column %)** | 3,627,000 (18.97%) | 47,000 (16.55%) | 47,000 (21.86%) | 10,000 (16.95%) | 15,000 (12.50%) | 3,778,000 (18.91%) |
| **Q4, N (column %)** | 4,049,000 (21.17%) | 57,000 (20.07%) | 29,000 (13.49%) | 7,000 (11.86%) | 14,000 (11.67%) | 4,184,000 (20.94%) |
| **Q5 (Highest), N (column %)** | 4,310,000 (22.54%) | 84,000 (29.58%) | 23,000 (10.70%) | 5,000 (8.47%) | 8,000 (6.67%) | 4,452,000 (22.28%) |

<sup>1</sup> Defined as 4 or more standard drinks for females during one occasion in the past 12 months, and 5 or more standard drinks for males during one occasion in the past 12 months
<sup>2</sup> Defined as any use in the past 12 months
<sup>3</sup> Defined as any use in the past 12 months
Note Data pooled from 2009 to 2014, total weighted sample size n=19,980,000 individuals

Table 2 shows the adjusted (i.e. model 1a-5a) and unadjusted (i.e. models 1b-5b) models for men. In fully adjusted models, while Gay men had 57% greater odds of reporting poor mental health (OR=1.57, 95% CI 1.15 to 2.16, p=0.005) relative to their heterosexual counterparts, there was no evidence that they had different odds of poor physical health, binge drinking, illicit drug use, or cannabis use. Bisexual men had 176% greater odds of reporting poor mental health (OR=2.76, 95% CI 2.01 to 3.77, p<0.001), and 57% greater odds of poor physical health (OR=1.57, 95% CI 1.10 to 2.25, p=0.013), but no evidence for differences in binge drinking, illicit drug use, or cannabis use. Men in the ‘don’t know’ group showed no elevated odds of poor mental or physical health; however, they had reduced odds of binge drinking by a factor of 0.35 (95% CI 0.19 to 0.62, p<0.001), reduced odds of illicit drug use by a factor of 0.16 (95% CI 0.08 to 0.32, p<0.001), and reduced odds of cannabis use by a factor of 0.16 (95% CI 0.08 to 0.30, p<0.001). Similarly, men in the ‘refuse’ group had no differences in mental or physical health, but had reduced odds of binge drinking (OR=0.40, 95% CI 0.24 to 0.66, p<0.001), illicit drug use (OR=0.43, 95% CI 0.22 to 0.86, p=0.016), and cannabis use (OR=0.38, 95% CI 0.19 to 0.80, p=0.009).

**Table 2.**
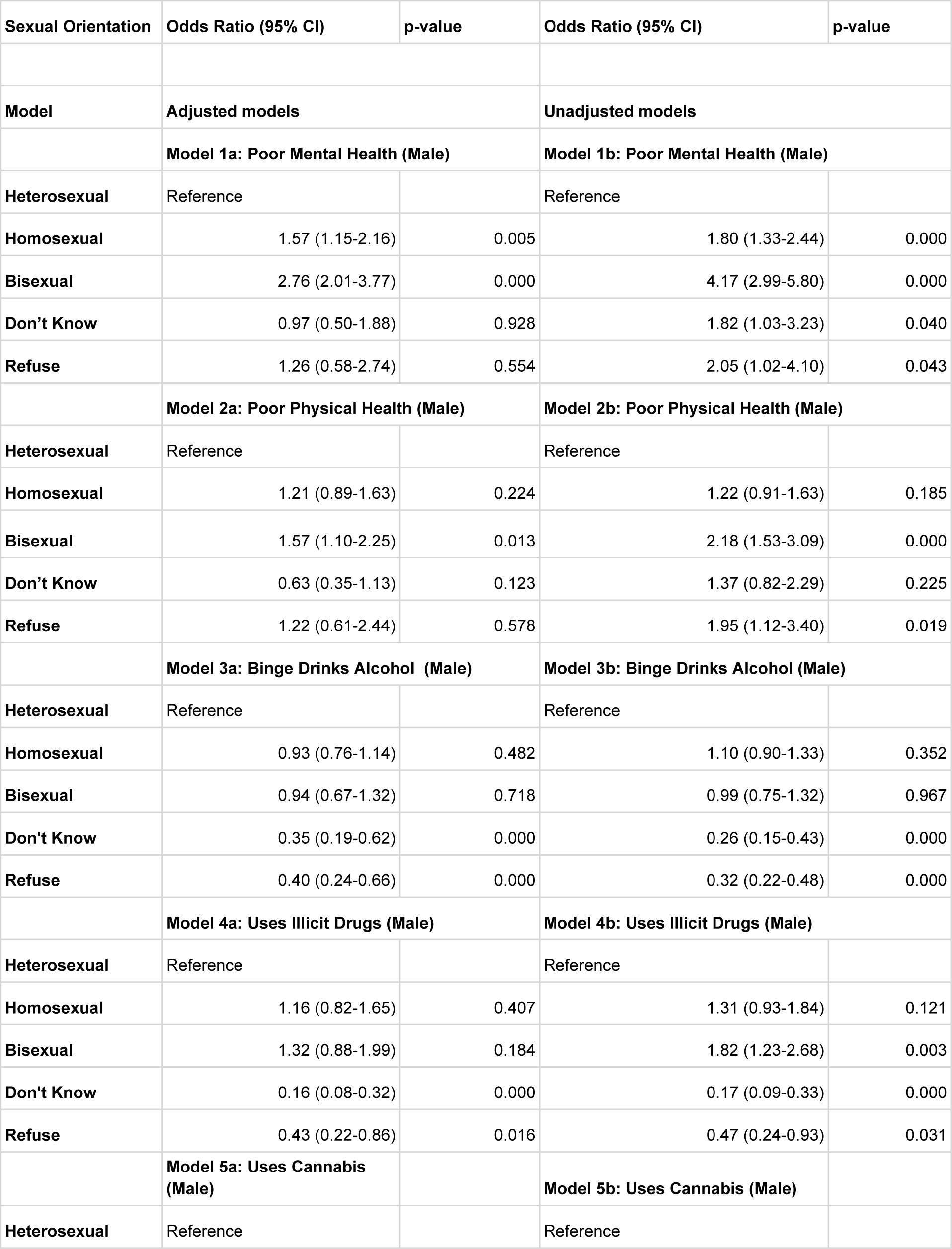
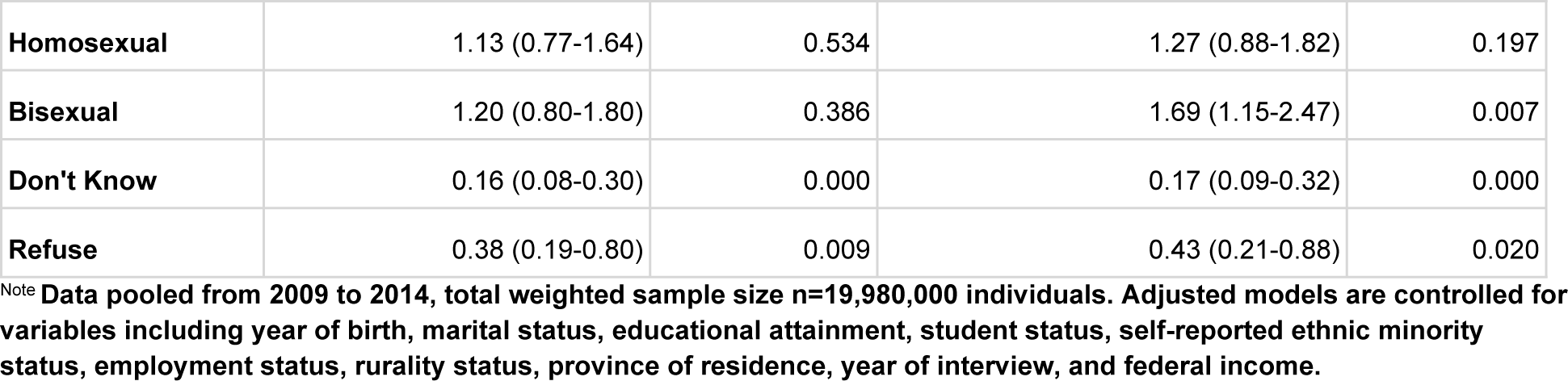
Male adjusted and unadjusted odds ratios for poor physical health, poor mental health, binge drinking, using illicit drugs, and using cannabis across sex and sexual orientation for Canadians.

| Sexual Orientation | Odds Ratio (95% CI) | p-value | Odds Ratio (95% CI) | p-value |
| --- | --- | --- | --- | --- |
| **Model** | **Adjusted models** |  | **Unadjusted models** |  |
|  | **Model 1a: Poor Mental Health (Male)** |  | **Model 1b: Poor Mental Health (Male)** |  |
| **Heterosexual** | Reference |  | Reference |  |
| **Homosexual** | 1.57 (1.15-2.16) | 0.005 | 1.80 (1.33-2.44) | 0.000 |
| **Bisexual** | 2.76 (2.01-3.77) | 0.000 | 4.17 (2.99-5.80) | 0.000 |
| **Don't Know** | 0.97 (0.50-1.88) | 0.928 | 1.82 (1.03-3.23) | 0.040 |
| **Refuse** | 1.26 (0.58-2.74) | 0.554 | 2.05 (1.02-4.10) | 0.043 |
|  | **Model 2a: Poor Physical Health (Male)** |  | **Model 2b: Poor Physical Health (Male)** |  |
| **Heterosexual** | Reference |  | Reference |  |
| **Homosexual** | 1.21 (0.89-1.63) | 0.224 | 1.22 (0.91-1.63) | 0.185 |
| **Bisexual** | 1.57 (1.10-2.25) | 0.013 | 2.18 (1.53-3.09) | 0.000 |
| **Don't Know** | 0.63 (0.35-1.13) | 0.123 | 1.37 (0.82-2.29) | 0.225 |
| **Refuse** | 1.22 (0.61-2.44) | 0.578 | 1.95 (1.12-3.40) | 0.019 |
|  | **Model 3a: Binge Drinks Alcohol (Male)** |  | **Model 3b: Binge Drinks Alcohol (Male)** |  |
| **Heterosexual** | Reference |  | Reference |  |
| **Homosexual** | 0.93 (0.76-1.14) | 0.482 | 1.10 (0.90-1.33) | 0.352 |
| **Bisexual** | 0.94 (0.67-1.32) | 0.718 | 0.99 (0.75-1.32) | 0.967 |
| **Don't Know** | 0.35 (0.19-0.62) | 0.000 | 0.26 (0.15-0.43) | 0.000 |
| **Refuse** | 0.40 (0.24-0.66) | 0.000 | 0.32 (0.22-0.48) | 0.000 |
|  | **Model 4a: Uses Illicit Drugs (Male)** |  | **Model 4b: Uses Illicit Drugs (Male)** |  |
| **Heterosexual** | Reference |  | Reference |  |
| **Homosexual** | 1.16 (0.82-1.65) | 0.407 | 1.31 (0.93-1.84) | 0.121 |
| **Bisexual** | 1.32 (0.88-1.99) | 0.184 | 1.82 (1.23-2.68) | 0.003 |
| **Don't Know** | 0.16 (0.08-0.32) | 0.000 | 0.17 (0.09-0.33) | 0.000 |
| **Refuse** | 0.43 (0.22-0.86) | 0.016 | 0.47 (0.24-0.93) | 0.031 |
|  | **Model 5a: Uses Cannabis (Male)** |  | **Model 5b: Uses Cannabis (Male)** |  |
| **Heterosexual** | Reference |  | Reference |  |
| **Homosexual** | 1.13 (0.77-1.64) | 0.534 | 1.27 (0.88-1.82) | 0.197 |
| **Bisexual** | 1.20 (0.80-1.80) | 0.386 | 1.69 (1.15-2.47) | 0.007 |
| **Don't Know** | 0.16 (0.08-0.30) | 0.000 | 0.17 (0.09-0.32) | 0.000 |
| **Refuse** | 0.38 (0.19-0.80) | 0.009 | 0.43 (0.21-0.88) | 0.020 |

Note Data pooled from 2009 to 2014, total weighted sample size n=19,980,000 individuals. Adjusted models are controlled for variables including year of birth, marital status, educational attainment, student status, self-reported ethnic minority status, employment status, rurality status, province of residence, year of interview, and federal income.

Table 3 shows the adjusted (i.e. model 1a-5a) and unadjusted (i.e. models 1b-5b) models for women where lesbians showed no evidence for a difference in poor mental or physical health relative to their heterosexual counterpart, but had 56% greater odds of binge drinking (95% CI 1.19 to 2.05, p=0.001), 83% greater odds of illicit drug use (95% CI 1.29 to 2.61, p=0.001), and 76% greater odds of cannabis use (95% CI 1.23 to 2.52, p=0.002). Relative to their heterosexual counterparts, Bisexual women exhibited the largest disparity for all health outcomes: 230% greater odds of poor mental health (95% CI 2.58 to 4.22, p<0.001), 139% greater odds of poor physical health (95% CI 1.84 to 3.10, p<0.001), 68% greater odds of binge drinking (95% CI 1.36 to 2.08, p<0.001), 134% greater odds of illicit drug use (95% CI 1.73 to 3.17, p<0.001), and 133% greater odds of cannabis use (95% CI 1.73 to 3.14, p<0.001). While women in the ‘don’t know’ group showed no evidence of differences in poor mental health, poor physical health, or cannabis use, they had lower odds of binge drinking (OR=0.35, 95% CI 0.22 to 0.55, p<0.001) and lower odds of illicit drug use (OR=0.28, 95% CI 0.08 to 0.96, p=0.043). Women in the ‘refuse’ group had no evidence of differences in poor mental health, poor physical health, Illicit drug use, or cannabis use, but had lower odds of binge drinking by a factor of 0.54 (95% CI 0.30 to 0.98, p=0.042). Generally, adjusting for covariates reduced the magnitude of associations between sexual orientation and health outcomes, but the patterns of significance— where the same groups exhibited significant differences in outcomes relative to heterosexuals— remained unchanged.

**Table 3.**
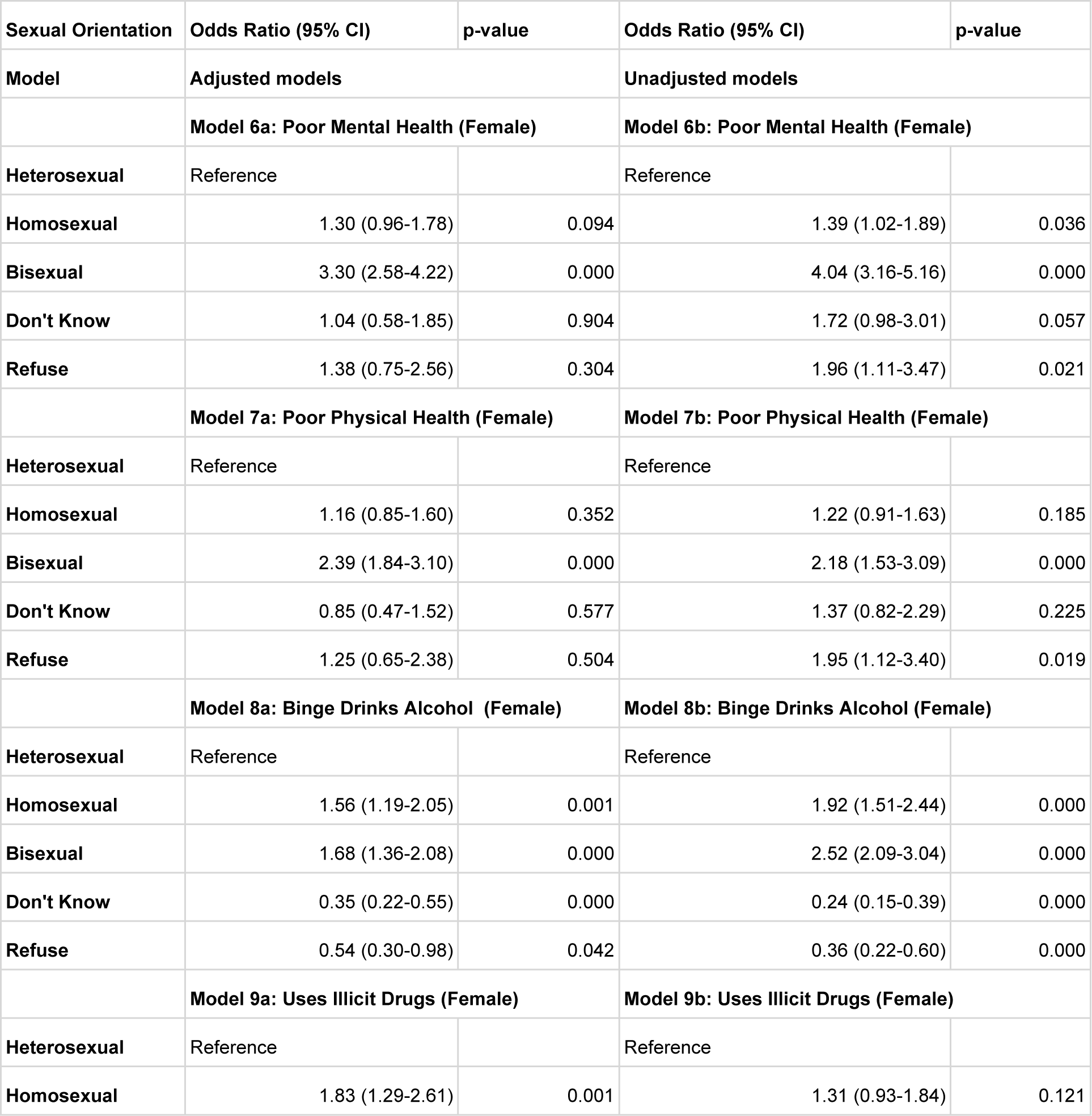
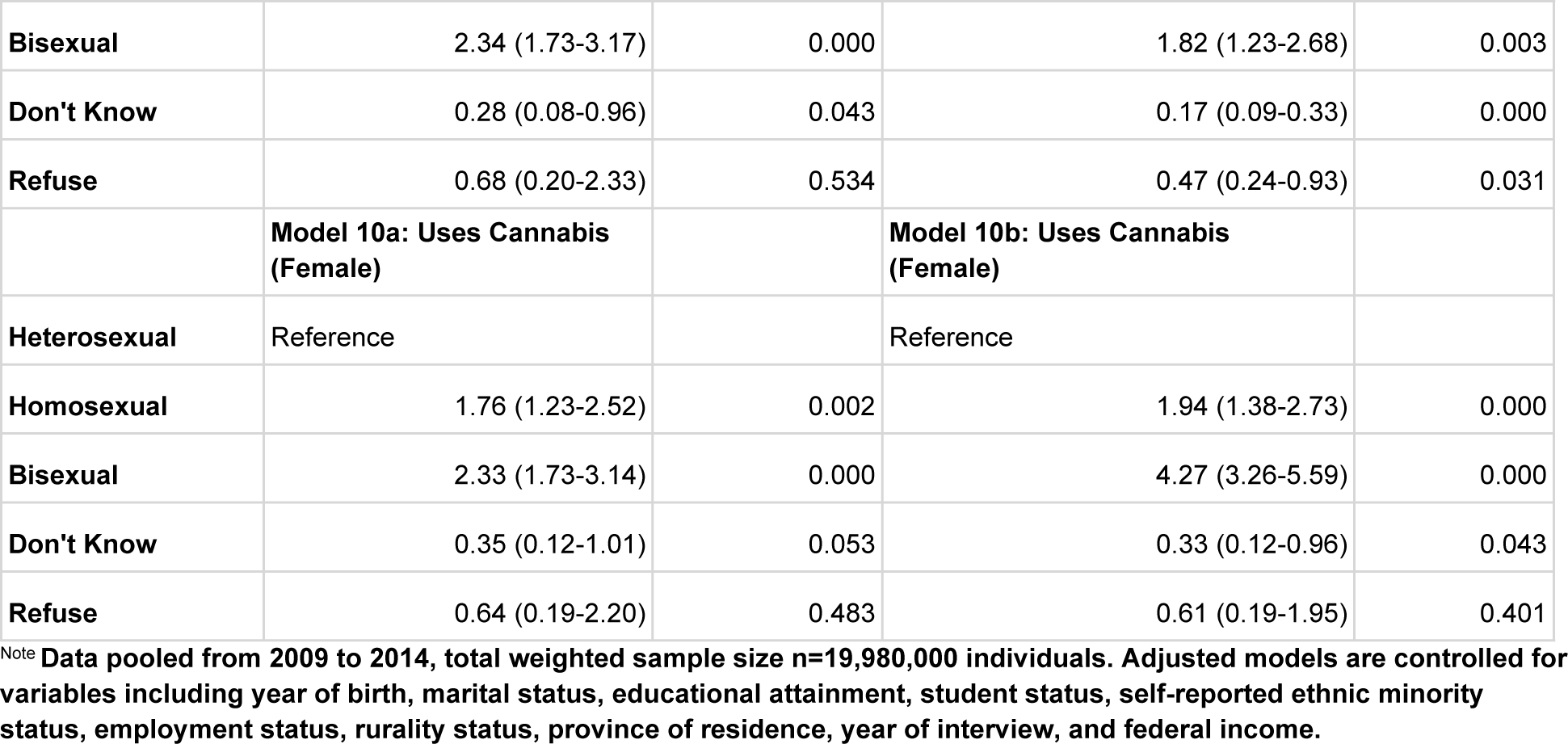
Female adjusted and unadjusted odds ratios for poor physical health, poor mental health, binge drinking, using illicit drugs, and using cannabis across sex and sexual orientation for Canadians.

| **Sexual Orientation** | **Odds Ratio (95% CI)** | **p-value** | **Odds Ratio (95% CI)** | **p-value** |
| --- | --- | --- | --- | --- |
| **Model** | **Adjusted models** |  | **Unadjusted models** |  |
|  | **Model 6a: Poor Mental Health (Female)** |  | **Model 6b: Poor Mental Health (Female)** |  |
| **Heterosexual** | Reference |  | Reference |  |
| **Homosexual** | 1.30 (0.96-1.78) | 0.094 | 1.39 (1.02-1.89) | 0.036 |
| **Bisexual** | 3.30 (2.58-4.22) | 0.000 | 4.04 (3.16-5.16) | 0.000 |
| **Don't Know** | 1.04 (0.58-1.85) | 0.904 | 1.72 (0.98-3.01) | 0.057 |
| **Refuse** | 1.38 (0.75-2.56) | 0.304 | 1.96 (1.11-3.47) | 0.021 |
|  | **Model 7a: Poor Physical Health (Female)** |  | **Model 7b: Poor Physical Health (Female)** |  |
| **Heterosexual** | Reference |  | Reference |  |
| **Homosexual** | 1.16 (0.85-1.60) | 0.352 | 1.22 (0.91-1.63) | 0.185 |
| **Bisexual** | 2.39 (1.84-3.10) | 0.000 | 2.18 (1.53-3.09) | 0.000 |
| **Don't Know** | 0.85 (0.47-1.52) | 0.577 | 1.37 (0.82-2.29) | 0.225 |
| **Refuse** | 1.25 (0.65-2.38) | 0.504 | 1.95 (1.12-3.40) | 0.019 |
|  | **Model 8a: Binge Drinks Alcohol (Female)** |  | **Model 8b: Binge Drinks Alcohol (Female)** |  |
| **Heterosexual** | Reference |  | Reference |  |
| **Homosexual** | 1.56 (1.19-2.05) | 0.001 | 1.92 (1.51-2.44) | 0.000 |
| **Bisexual** | 1.68 (1.36-2.08) | 0.000 | 2.52 (2.09-3.04) | 0.000 |
| **Don't Know** | 0.35 (0.22-0.55) | 0.000 | 0.24 (0.15-0.39) | 0.000 |
| **Refuse** | 0.54 (0.30-0.98) | 0.042 | 0.36 (0.22-0.60) | 0.000 |
|  | **Model 9a: Uses Illicit Drugs (Female)** |  | **Model 9b: Uses Illicit Drugs (Female)** |  |
| **Heterosexual** | Reference |  | Reference |  |
| **Homosexual** | 1.83 (1.29-2.61) | 0.001 | 1.31 (0.93-1.84) | 0.121 |
| **Bisexual** | 2.34 (1.73-3.17) | 0.000 | 1.82 (1.23-2.68) | 0.003 |
| **Don't Know** | 0.28 (0.08-0.96) | 0.043 | 0.17 (0.09-0.33) | 0.000 |
| **Refuse** | 0.68 (0.20-2.33) | 0.534 | 0.47 (0.24-0.93) | 0.031 |
|  | **Model 10a: Uses Cannabis (Female)** |  | **Model 10b: Uses Cannabis (Female)** |  |
| **Heterosexual** | Reference |  | Reference |  |
| **Homosexual** | 1.76 (1.23-2.52) | 0.002 | 1.94 (1.38-2.73) | 0.000 |
| **Bisexual** | 2.33 (1.73-3.14) | 0.000 | 4.27 (3.26-5.59) | 0.000 |
| **Don't Know** | 0.35 (0.12-1.01) | 0.053 | 0.33 (0.12-0.96) | 0.043 |
| **Refuse** | 0.64 (0.19-2.20) | 0.483 | 0.61 (0.19-1.95) | 0.401 |

Note Data pooled from 2009 to 2014, total weighted sample size n=19,980,000 individuals. Adjusted models are controlled for variables including year of birth, marital status, educational attainment, student status, self-reported ethnic minority status, employment status, rurality status, province of residence, year of interview, and federal income.

The 3 sensitivity test results are overall consistent with the findings from main models. To begin with, the modified Poisson regression analyses (see Appendix, Table S3 models 1c-5c for males and Appendix, Table S4 models 6c-10c for females) confirmed these patterns of significance, showing consistent differences for the same groups when compared to their heterosexual counterparts. Second, models using complete case analyses (Appendix, Table S3 models 1d-5d for male, Appendix, Table S4 models 6d-10d for female) showed similar directionality, strength of association, and patterns of significant results compared to the imputed main analyses. Weighted number of item non-response for all variables can be found in Appendix Table S2 (weighted n=19,980,000), where level of missingness varied from 0% for variables such as student status and rurality to the highest at 4.49% for reports of income quintile. The third sensitivity test showed the trends of disparities (between each sexual orientation group vs. heterosexual) over time (Appendix Figure S1-S10), based on the sexual orientation interaction with ‘year of interview’ are shown in the linear trends are tested using the Mann-Kendall Trend Test (Appendix Table S1). Bisexual women were the only group to show evidence for an increasing disparity in mental health over our study period from 2009-2014 (tau=0.867, p=0.024). Evidence for a decreasing disparity in substance use was found across multiple groups including 1) gay mens’ illicit drug use (tau=-0.867, p=0.024) and cannabis use (tau=-0.867, p=0.024), 2) bisexual mens’ illicit drug use (tau=-0.867, p=0.024) and cannabis use (tau=-0.867, p=0.024), as well as 3) lesbian womens illicit drug use (tau=-0.867, p=0.024).

## Discussion

Our study provides details of health disparities in a broad set of health outcomes across sexual orientation using a population-representative sample of Canada. Among LGB participants, there were evidence for elevated risk of poor mental health (i.e. gay men, bisexual men, bisexual women), poor physical health (i.e. bisexual men, bisexual women), binge drinking (i.e. lesbians, bisexual women), illicit drug use (i.e. lesbians, bisexual women), and cannabis use (i.e. lesbians, bisexual women) relative to their heterosexual counterparts. While prior studies have generally highlighted worse health outcomes among sexual minorities, our study provides evidence that the level and number of health disparities vary among sexual minority groups: from gay men showing elevated risk in 1 outcome tested (i.e. poor mental health) to bisexual women who had elevated risk across all 5 outcomes tested. Furthermore, the strength of disparity varies widely between groups, e.g. with lesbian having 76% increased odds of cannabis use relative to heterosexual women, while bisexual women having 133% increased odds.

### Substance use in sexual minority women

A previous study examining the association between sexual orientation and substance use hospitalisation across Canada (also based on the Canadian community health survey) found that while bisexual women had elevated odds of substance use hospitalizations by a factor of 2.46 compared to their heterosexual counterpart [44], no elevated risk was detected in other sexual minority groups. Given that bisexual women had elevated risk in each substance use outcome in our study, and had the largest disparities relative to their heterosexual counterparts (among all other LGB groups), it was not surprising to see that this group also had heightened risk for substance-related hospitalisation. On the other hand, lesbians in our study also exhibited increased risk for all 3 substance use outcomes (with a smaller effect size compared to bisexual women); however, there was no evidence in the prior study that the lesbian group had higher risk of hospitalizations for any substances (Hazard Ratio, HR=0.98, 95% CI 0.47 to 2.06), alcohol-related hospitalizations (HR=0.98, 95% CI 0.34 to 2.86), or illicit drugs and cannabis hospitalizations (HR=0.96, 95% 0.38 to 2.39). While both lesbians and bisexual women report high levels of substance use in our study, only bisexual women have a higher risk of hospitalization. This discrepancy is aligned with findings from a review indicating that bisexual identity is often associated with more problematic substance use than those identifying as strictly heterosexual or homosexual [45]. This literature helps to explain why, among women, despite elevated risks of substance use in lesbians and bisexual women, only the bisexual group was linked to increased substance use hospitalizations.

### Substance use in ‘don’t know’ and ‘refuse’ individuals

While all participants in the ‘refused’ and ‘don’t know’ had even odds of poor physical and mental health relative to their heterosexual counterparts, many reported significantly reduced odds of substance use (i.e. binge drinking, illicit drugs, and cannabis) including ‘don’t know’ men (all 3 substance outcomes), ‘refused’ men (3 substance outcomes), ‘don’t know’ women (2 outcomes), ‘refused’ women (1 outcome). These results starkly contrast with a prior study that examined substance-related hospitalisation across sexual orientation [44]. More specifically, it showed that the ‘don’t know’ and ‘refused’ groups (combined into a single ‘other’ group) had elevated risk of age-adjusted incidence rates for substance-use hospitalizations [44]. In fully adjusted models, no differences were shown between the ‘other’ groups and their gender matched heterosexual counterparts for all substances examined. The discrepancy between self-reported substance use and substance use-related hospitalisation rates among the ‘don’t know’ and ‘refused’ groups raises questions about the accuracy of self-reports in these groups. Further research should investigate whether social desirability bias or fear of stigma associated with substance use may affect self-reported data in groups that are also reluctant to disclose their sexual orientations.

### Mental and physical health in sexual minority individuals

Among LGB men and women, all groups, except for lesbians, showed elevated odds of reporting poor mental health. A prior literature review has shown that, in most studies, LGB individuals report higher risk of mental health problems [46]. However, in a controlled sibling study, involving comparison of lesbians to their heterosexual sisters found no statistically significant differences in mental health, and even found lesbians to have higher rates of self esteem [47]. The author hypothesised that for lesbians, their degree of disclosure about their sexual orientation, or ‘outness’, may have acted as a protective factor in their study participants. Our study, based on self-reported mental health, also provides some evidence that lesbians may uniquely be protected against poor mental health among sexual minority groups.

Our study shows that while gay and lesbian individuals had similar risk of poor physical health as their heterosexual counterparts, bisexual men and women had higher risk of poor physical health. Prior studies on differences in physical health across sexual orientations had different results, which may be due to differences in how gender and sexual orientation are disaggregated. For example, a recent study using the US General Social Survey [48], found that after controlling for sociodemographic variables, no LGB groups have significant differences in their physical health over the past 30 days relative to their heterosexual counterparts; however, the study did not have gender disaggregated analyses, or used sex-interactions, to explore sex/gender effect modification (leaving the possibility that a bisexual vs homosexual physical health difference may exist in 1 sex and not the other in their sample). In a Swedish nationally representative study [7], after statistical adjustments, LGB individuals reported higher odds of physical symptoms (e.g., pain, insomnia, dermatitis, tinnitus, intestinal problems) and conditions (e.g., diabetes, asthma, high blood pressure) compared to heterosexuals; however, the study did not disaggregate homosexual and bisexual participants, and studied sexual minority as a single group, which provided limited insights into the bisexual vs gay/lesbian differences that we found.

Our finding that bisexual individuals (and not gay men/lesbians) had elevated risk of poor physical health may be explained by increased minority stress as past studies have indicated that biphobia is pervasive in heterosexual and LGBTQ+ communities [30,49], and as a result, bisexual individuals receive reduced community support and face a higher level of discrimination compared to gay/lesbian individuals [50]. In a systematic review [6], minority stress has been linked to poor physical health through direct physiological stress response (e.g. immune dysregulation and allostatic load) and can modify health behaviours through distress/psychopathology. Given the elevated levels of minority stress in bisexual individuals, it is unsurprising that both mental and physical health disparities are heightented in bisexual individuals.

### Changes in disparities over time (2009-2014)

The analysis of time trends has shed light on the changing dynamics of health outcomes over time and the disparities faced by various sexual orientation groups. These trends highlight the evolving nature of health disparities, though it is important to consider the potential limitations in interpreting these changes due to shifts in population composition across multiple cross-sectional periods. As such, these findings on temporal changes warrant careful consideration. Key observations include: 1) An increasing disparity in mental health issues among bisexual women, which raises concerns given their already higher vulnerability to adverse health outcomes; 2) A significant decrease in substance use disparities among several sexual orientation groups, notably gay and bisexual men for illicit drug and cannabis use, and lesbians for illicit drug use. This decline in substance use among these groups is encouraging. However, the potential widening gap in mental health disparities among bisexual women calls for urgent, focused intervention strategies.

### Limitations and strengths

There are a number of limitations in our study. First, while the study included sex, there were no measures of gender identity, and we were unable to identify health disparities in gender diverse groups including transgender and non-binary individuals. Second, sexual orientation were based on self-identification, but prior research has shown that a more comprehensive representation should include measures of attraction and sexual behaviour, with evidence showing that certain groups, including heterosexually identified men who have sex with men [51], may have even worse health outcomes compared to gay and bisexual identified men. Third, the study covered multiple years, and the levels of disparity across groups may have changed over these years. To help partially mitigate this concern, sensitivity tests were conducted to investigate the interaction between sexual orientation and time; however, since the data is cross-sectional (and not longitudinal), the observed changes may be driven by compositional change rather than reflecting a larger societal shift. Fourth, some groups might not openly share their substance use problems, casting doubt on the trustworthiness of their self-reports. Future studies should explore how social pressure or fear of stigma related to substance use could impact self-reported data in these groups, such as those who did not reveal their sexual orientations. Fifth, self-reported health measures may over- or underestimate true health status due to issues like social desirability bias, recall bias, and the inability of participants to accurately quantify their health experiences [52]. Sixth, the study utilized data from 2009-2014, and it is possible that the levels of disparity across groups have evolved since then. While this timeframe may limit the current applicability of our findings, the analyses remain valuable by providing a historical baseline against which to measure future shifts in health disparities. Finally, while the control variables were chosen based on prior literature, there may still be uncontrolled confounders leading to residual confounding.

Despite these limitations, our study has several notable strengths. First, we used a range of health outcomes to capture multiple dimensions of health inequities. By examining various self-rated health outcomes, ranging from physical and mental health to substance use, the study provides a more comprehensive picture of health disparities across sexual orientations in Canada to help inform health promotion strategies. Second, the use of a nationally representative sample is a strength of this study. This was supplemented by the use of population weights provided by Statistics Canada to adjust for potential discrepancies in response rates compared to characteristics found in the Canadian Census, which may improve the generalizability of our study to the Canadian population. Third, the use of disaggregated data for sex and sexual orientation helped to identify significant differences between sexual minority groups (i.e. increased risk of all health outcomes shown in bisexual women), which helps to highlight specific sexual minority subgroups for targeted interventions.

## Conclusions

This study highlights significant health disparities among sexual minorities in Canada, particularly emphasising the unique challenges faced by bisexual women. The findings suggest an urgent need for tailored interventions, as bisexual women not only report higher levels of substance use but also face greater health risks, including hospitalization. A key concern highlighted is the persistence of biphobia and the need for interventions to specifically address this, such as making resources on biphobia readily available in clinical and community settings and ensuring frontline workers are equipped to handle crises related to bisexual stigma. The study also underlines the need for cautious interpretation of self-reported data, especially from individuals uncertain or uncomfortable with disclosing their sexual orientation. Discrepancies noted in self-reported substance use versus administrative data underscore the necessity for further research in clinical settings to ensure accurate health assessments and interventions. Our repeated cross-sectional analysis exposes persistent and sometimes worsening disparities across sexual orientations over time, underscoring the need for future research to adopt longitudinal approaches. Such research would offer insights into evolving health challenges and the effectiveness of past interventions, helping to shape targeted prevention strategies.

In conclusion, this research, utilising a nationally representative sample and comprehensive analyses, provides insights into the complex landscape of health inequities faced by the LGB community. The heightened vulnerability of bisexual women across all assessed health metrics calls for immediate and specific public health responses. Future efforts must continue to refine these interventions and assess their impact longitudinally to ensure they are culturally appropriate and truly effective.

## Data Availability

Third party data used in this study is owned by and is considered confidential by Statistics Canada. It can be accessed for research purposes through the Canadian Research Data Centre Network (https://crdcn.org). The data can be accessed in secure computing environments (inside designated Research Data Centres) upon approval by Statistics Canada of a project proposal. CCHS data is available through the Research Data Centres program administered by Statistics Canada (see link for eligibility and process to request access: https://www.statcan.gc.ca/eng/rdc/index) for researchers who meet the criteria for access to confidential data.

